# Short-term Functional Outcomes of Patients with acute intracerebral hemorrhage in the Native and Expatriate Population

**DOI:** 10.1101/2023.10.17.23297178

**Authors:** Naveed Akhtar, Mahesh Kate, Saadat Kamran, Sujatha Joseph, Deborah Morgan, Ryan Uy, Blessy Babu, Shobhna Shanti, Ashfaq Shuaib

## Abstract

**Objectives:** Functional outcomes in patients with intracerebral hemorrhage (ICH) have not been well characterized in the Middle East and North Africa Region. We report the 30 and 90-day clinical outcomes in the native and expatriate of Qatar with ICH.

**Methods:** We evaluated the Glasgow Coma Scale (GCS), NIHSS, and imaging in the Qatar Stroke Registry (2013–22). The outcome measures were a modified Rankin Scale (mRS) at 90 days and mortality at 30 and 90 days. Unfavourable outcome was defined as mRS of 4-6. We performed non-parametric ROC analyses to measure the concordance index (C-index) to assess the goodness-of-fit of ICH score for predicting 30 day and 90-day mortality and functional outcome.

**Results:** 1660 patients (median age of 49 (41.5-58) years; male 83.1%, expatriates 77.5%) with ICH, including supratentorial deep in 65.2%, cortical in 16.2%, infratentorial 16% and primary intraventricular in 2.5% were studied. The median baseline ICH volume was 7.5 (3.2-15.8) ml. An unfavorable outcome was seen in 673 (40.5%) patients at 90 days. Mortality at 30 days and 90 days was 10.4% and 15.1%. Increasing age (OR (95% CI), 1.02 (1.00-1.03)), lower GCS (0.77 (0.73-0.80)), prior use of antiplatelet medications (1.82 (1.19-2.08)), higher ICH volume (1.03 (1.02-1.04)), and presence of any intraventricular hemorrhage (1.57(1.19-2.08)), were associated with unfavorable outcome.

**Conclusions:** In this relatively younger ICH cohort more than 75% were expatriates, had smaller ICH volume and had better functional outcomes. Prognostic scoring systems may have to be modified in this population to avoid early withdrawal of care.

## Introduction

Despite improvements in medical and surgical therapies, acute intracerebral hemorrhage (ICH) carries a grave prognosis (1), with a 30-day mortality between 30-55% and fewer than 20% patients are functionally independent 6 months (2–5). Several tools are used to predict mortality and functional outcome following an ICH (6–11) with the ICH-score being one of the most common in clinical practice (6). A recent meta-analysis of 55 studies reported that ICH-score, while having a good discrimination with low scores, overestimated mortality with higher scores (12).

The clinical impact of ICH appears disproportionately high among lower-resource populations (13). In US-based studies, ICH incidence has been reported to be ≈1.6-fold greater among Black and Mexican (13). Internationally, ICH incidence is substantially higher in low-income countries and comprises ≈ 18% of all strokes (2–4). There are a few reports outlining prognosis of ICH from low resource countries (14-25]). The number of patients in such series are small 19,20,21,22,24), and the information collected retrospectively (18,22,23) making prediction modelling difficult to study.

In order to study admission trends, risk factors and prognosis in a large multiethnic population of patients from native (Arabs), and expatriates (South Asian, Southeast Asian), we investigated primary ICH admissions in the tertiary care centre in Qatar. We aimed to evaluate the outcome and prognosis in patients with ICH.

## Methods

The Qatar Stroke Database was established in March 2013 at the Hamad General Hospital (HGH), Doha. All data on patients with suspected acute stroke, including transient ischemic attacks (TIAs), cerebral infarction, stroke mimics, ICH and cerebral venous thrombosis (CVT) are entered prospectively into the registry (26–30). Depending on the severity and location of the ICH, patients are admitted to the stroke ward, ICU or neurosurgery.

For inclusion in the study, the following criteria were use: A diagnosis of ICH that was proven by imaging and related to hypertension or amyloid angiopathy. Patients with primary ventricular ICH were included but hemorrhages related to AVMs or aneurysms were excluded. We also excluded ICH related to trauma, hemorrhagic transformation of an ischemic infarction and systemic coagulopathy. Patients with primary subdural hemorrhage and subarachnoid hemorrhage were also excluded.

### Patient characteristics

Patient characteristics including age, sex, nationality, medical comorbidities, and prior medication were recorded. Ethnicity was defined based on social groups sharing cultural and language traits within the broad categories of Native Residents (Qatari and non-Qatari Arabs), South Asians (Indian, Pakistani, Bangladeshi, Sri Lankans, Nepalese and Myanmar) and the Southeast Asians (predominantly from Philippines). Data collected included pre-stroke modified Rankin scale (mRS), NIH Stroke Scale (NIHSS) score at admission, length of stay (LoS) in ED, LoS in the stroke ward (SW), general medical ward (GMW), neurosurgical ward or the ICH.

### Radiological variables

Patients’ plain head CT scans were analyzed to identify the following data: location (cortical, basal ganglia, thalamus, brainstem or cerebellum), ICH volume (cm3) measured using the method with the largest length in three dimensions divided by two (equation: ABC/2); presence or absence of intraventricular hemorrhage IVH).

### Clinical scores and Outcome Measures

All patients had Glasgow Coma Scale (GCS) measured at admission and regularly during admission. The primary outcome measures were unfavorable mRS (4–6) at 90-day, 30-day and 90-day mortality. The ICH-score was used to measure prognosis at admission. The ICH Score was the sum of individual points assigned as follows: GCS score 3 to 4 (=2 points), 5 to 12 (=1), 13 to 15 (=0); age 80-years yes (=1), no (=0); infratentorial origin yes (=1), no (=0); ICH volume >30 cm3 (=1), <30 cm3 (=0); and intraventricular hemorrhage yes (=1), no (=0) (6).

### Statistics

Continuous variables (age, systolic BP, diastolic BP, NIHSS, GCS, BMI and ICH volume) are presented as median values with interquartile range. Categorical variables (sex, residential status, mode of arrival to the hospital, medical comorbidities, concomitant medications, hemorrhage location, ICH score, hospital admission status, and discharge disposition) are presented as counts and percentages. The study population was divided in two groups patients with favorable (mRS 0-3) and unfavorable (mRS 4-6) 90-day outcomes. Unadjusted univariable binary logistic regression analyses were performed to assess differences in continuous variables. The chi-square test was employed to compare categorical variables between groups. Variables with a p-value <0.05 were used for regression analyses. Multivariable binary logistic regression analyses were used to assess the effects of the individual predictors with respect to 30-day, 90-day mortality and 90-day functional outcome. We also performed non-parametric receiver operator curve (ROC) analyses to measure the area under the curve (AUC) for all the variables used in the multivariable logistic regression analysis. We performed non-parametric ROC analyses to measure the concordance index (C-index) to assess the goodness-of-fit of ICH score for predicting 30 day, 90-day mortality and functional outcome at 90 days. All statistical analysis was performed using STATA 18.0 BE (StatCorp LLC Texas, USA).

The study was approved by the Committee for Human Ethics Research, Academic Health Service at HMC (MRC-01-18-102).

## Results

### Overall results

1660 patients with primary ICH were available for analysis. As shown in Figure-1, ICH represented 14.8% (1674/11338) patients with final diagnosis of stroke. The median (IQR) age was 49 (41.5-58) years, men were (1379, 83.1%) and expatriates were (1286, 77.5%).

**Figure 1:**
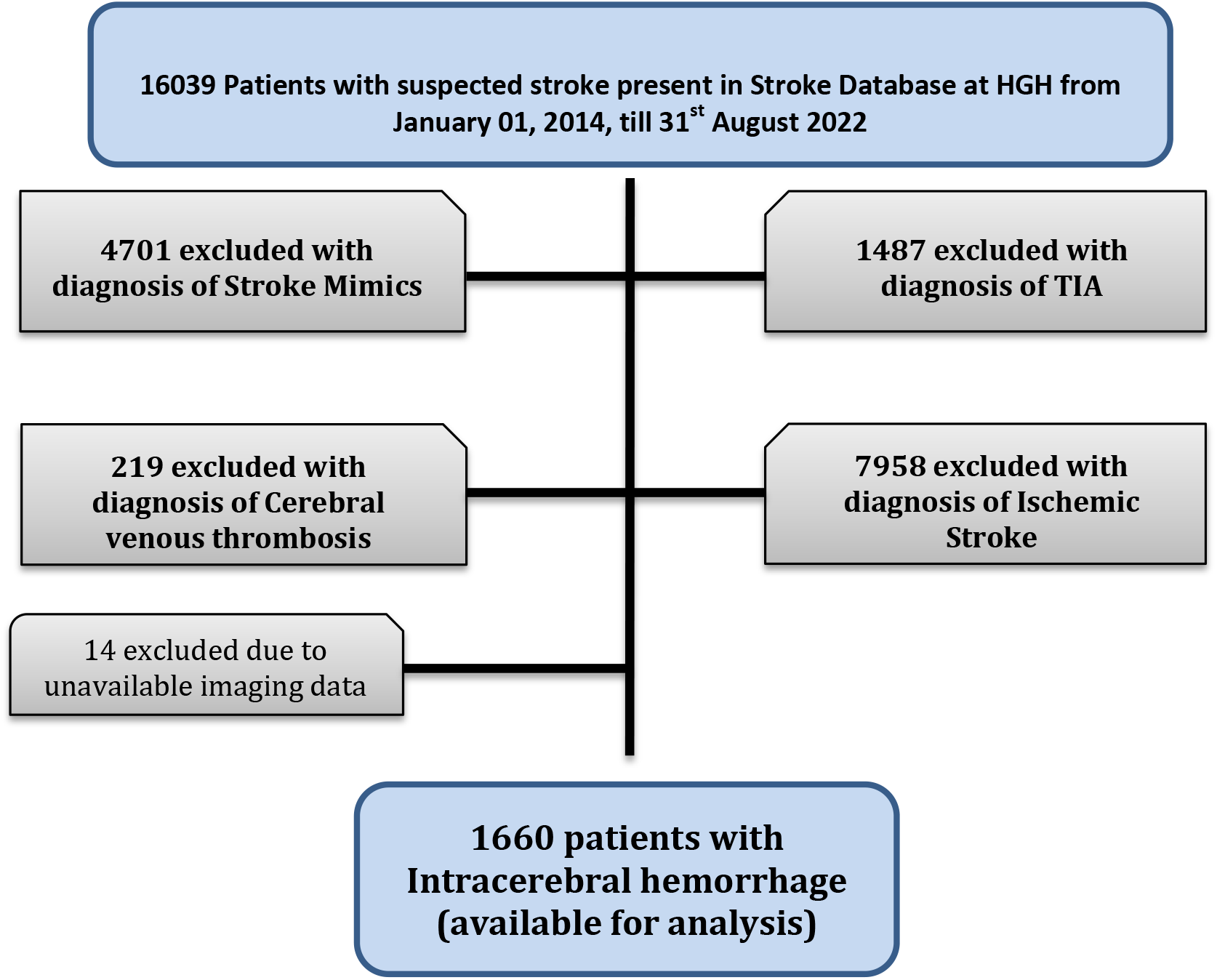
Study population for the study.

The details of the clinical features, risk factors, imaging, NIHSS and GCS at admission, ICH score, location and volume, admission location, and surgical intervention are summarized in Table-1. We compared the variables in two groups: favorable outcome (mRS 0-3) versus unfavorable outcome (mRS 4-6) at 90 days. A favorable outcome was seen in 987 (59.5%). The better outcome in men compared to women may be related to the relatively young age of the male (48, 41-57 years) patients compared to female (53, 44-66 years, OR 0.96 95%CI 0.95-0.97) (Figure-2). The older women reflect the higher percentage of native residents (125, 33.4%) compared to expatriate women (156, 12.1%, OR 3.6 95%CI 2.7-4.8) in the study population.

**Figure 2:**
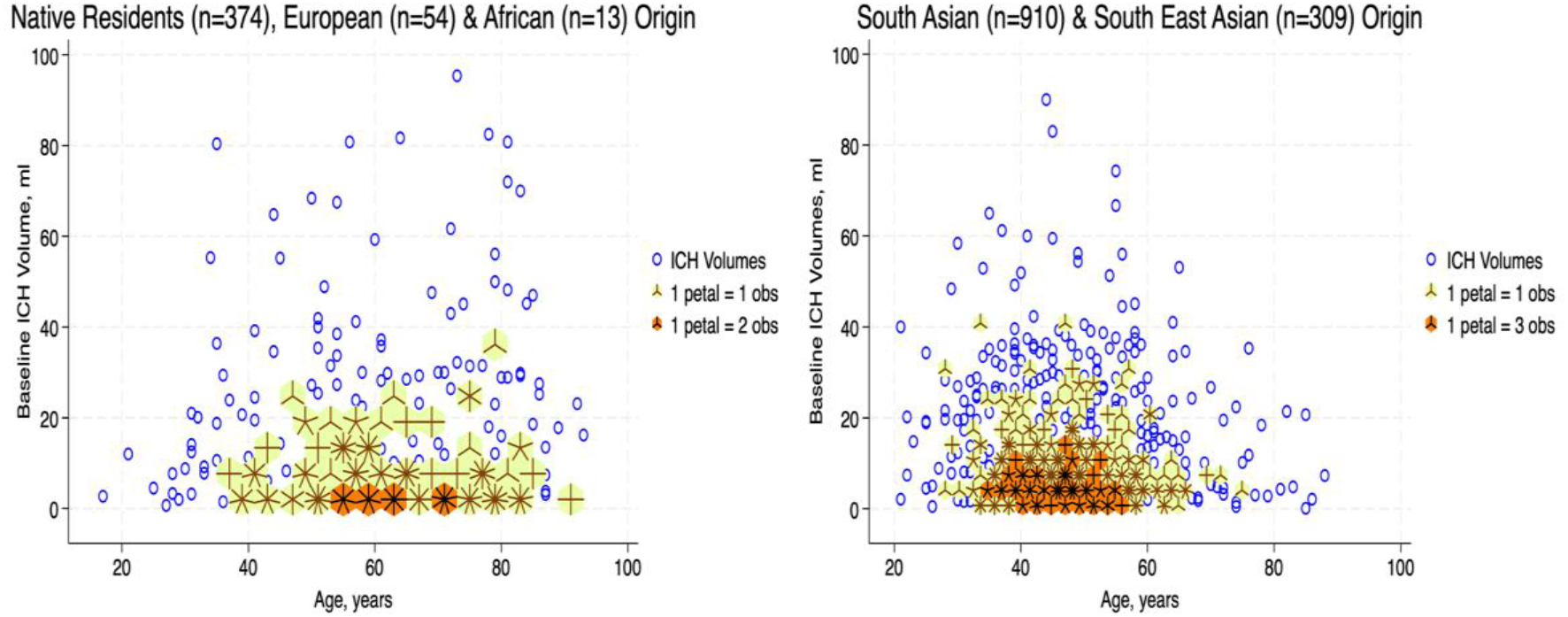
Density-Distribution Sunflower Plot of baseline intracerebral volume, ml and age. ICH patients who were Native residents, European and African Origin were older compared to the patients of South Asian and South East Asian Origin.

**Table 1.** Clinical, imaging, hospital admission and outcome characteristics in the favourable (modified Rankin scale 0-3) outcome at 90 days group and unfavourable (modified Rankin scale 4-6) at 90 days outcome group.

| Variable | Favorable Prognosis (n=987, 59.5) | Unfavorable Prognosis (n=673, 40.5) | Total (n=1660) | p value |
| --- | --- | --- | --- | --- |
| **Demographics** |  |  |  |  |
| Median (IQR) Age, years | 48 (41-56) | 50 (42-61) | 49 (41.5-58) | <0.001 |
| Female, n (%) | 149 (9) | 132 (7.9) | 281 (16.9) | 0.016 |
| Male, n (%) | 838 (50.5) | 541 (32.6) | 1379 (83.1) |  |
| Native Residents n (%) | 190 (11.4) | 184 (11.1) | 374 (22.5) | <0.001 |
| **Expatriate** |  |  |  |  |
| South Asian n (%) | 555(33.4) | 355(21.4) | 910 (54.8) |  |
| Southeastern Asian n (%) | 200 (12.1) | 109 (6.6) | 309 (18.6) |  |
| African n (%) | 30 (1.8) | 24 (1.4) | 54 (3.3) |  |
| North American/European n (%) | 12 (0.7) | 1 (0.1) | 13 (0.8) |  |
| **Modes of Arrival** |  |  |  |  |
| Emergency Medical System n (%) | 797 (48) | 587 (35.4) | 1384 (83.4) | <0.001 |
| Interfacility Transfer n (%) | 53 (3.2) | 44 (2.6) | 97 (5.8) |  |
| In-hospital Stroke n (%) | 6 (0.4) | 5 (0.3) | 11 (0.7) |  |
| Private Transport n (%) | 131 (7.9) | 37 (2.2) | 168 (10.1) |  |
| **Clinical Assessments** |  |  |  |  |
| Median Systolic blood pressure mmHg | 179 (154-204) | 182 (152-210) | 180 (153-206) | 0.096 |
| Median Diastolic blood pressure mmHg | 105 (90-121) | 106 (89-123) | 105.5 (90-122) | 0.827 |
| High Blood Pressure on Admission n (%) | 833 (50.2) | 550 (33.1) | 1383 (83.3) | 0.151 |
| Median Baseline NIHSS | 6 (2-12) | 20 (13-25) | 11 (4-20) | <0.001 |
| Median Glasgow Coma Scale | 15 (14-15) | 10 (6-14) | 14 (10-15) | <0.001 |
| GCS Score 13-15 | 844 (50.8) | 258 (15.5) | 1102 (66.4) | <0.001 |
| GCS Score 5-12 | 131 (7.9) | 287 (17.3) | 418 (25.2) |  |
| GCS Score 3-4 | 12 (0.7) | 128 (7.7) | 140 (8.4) |  |
| Median Baseline Blood Glucose, mg% | 7.2 (5.9-9.1) | 8.3 (6.8-11) | 7.5 (6.2-9.9) | <0.001 |
| Median BMI* Kg/m<sup>2</sup> | 27 (24.2-30) | 26.7 (24.2-29.7) | 27 (24.2-29.9) | 0.7 |
| **Medical Comorbidities** |  |  |  |  |
| Hypertension n (%) | 630 (38) | 425 (25.6) | 1055 (63.6) | 0.280 |
| Diabetes n (%) | 324 (19.5) | 241 (14.5) | 565 (34) | 0.208 |
| Dyslipidemia n (%) | 300 (18.1) | 165 (9.9) | 465 (28) | 0.009 |
| Prior Stroke n (%) | 47 (2.8) | 66 (4) | 113 (6.8) | <0.001 |
| Atrial Fibrillation n (%) | 21 (1.3) | 34 (2.1) | 55 (3.3) | 0.001 |
| Coronary Artery Disease n (%) | 45 (2.7) | 50 (3) | 95 (5.7) | 0.013 |
| Smoking n (%) | 124 (7.5) | 48 (2.9) | 172 (10.4) | <0.001 |
| **Concomitant Medications** |  |  |  |  |
| Antihypertensive use n (%) | 218 (13.1) | 176 (10.6) | 394 (23.7) | 0.056 |
| Anticoagulants use n (%) | 19 (1.1) | 30 (1.8) | 49 (2.9) | 0.003 |
| Antiplatelet use n (%) | 85 (5.1) | 105 (6.3) | 190 (11.4) | <0.001 |
| Antidiabetic use n (%) | 99 (6) | 91 (5.5) | 190 (11.5) | 0.028 |
| **Hemorrhage location** |  |  |  |  |
| Supratentorial Deep n (%) | 654 (39.4) | 429 (25.8) | 1083 (65.2) | <0.001 |
| Supratentorial Cortical n (%) | 174 (10.5) | 95 (5.7) | 269 (16.2) |  |
| Infratentorial n (%) | 127 (7.6) | 139 (8.4) | 266 (16) |  |
| Primary Intraventricular n (%) | 32 (1.9) | 10 (0.6) | 42 (2.5) |  |
| Any intraventricular n (%) | 243 (14.6) | 334 (20.1) | 577 (34.8) | <0.001 |
| Median Intraparenchymal Volume**, ml | 5.2 (2.5-10.5) | 13.2(5.8-24.4) | 7.5 (3.2-15.8) | <0.001 |
| Hemorrhage vol. ≥30 ml** | 30 (2) | 95 (6.3) | 125 (8.3) | <0.001 |
| **ICH score 0\*\*** | 524 (34.9) | 125 (8.3) | 649 (43.2) | <0.001 |
| 1 | 293 (19.5) | 167 (11.1) | 460 (30.7) |  |
| 2 | 63 (4.2) | 161 (10.7) | 224 (14.9) |  |
| 3 | 15 (1) | 108 (7.2) | 123 (8.2) |  |
| 4 | 2 (0.13) | 39 (2.6) | 41 (2.7) |  |
| 5 | 0 (0) | 4 (0.3) | 4 (0.3) |  |
| Stroke Unit care n (%) | 595 (35.8) | 131 (7.9) | 726 (43.7) | <0.001 |
| Intensive Unit Care n (%) | 357 (21.5) | 483 (29.1) | 840 (50.6) | <0.001 |
| Mechanical ventilation n (%) | 132 (8) | 432 (26) | 564 (34) | <0.001 |
| Extra-ventricular Drainage n (%) | 40 (2.4) | 78 (4.7) | 118 (7.1) | <0.001 |
| Craniotomy/Craniectomy n (%) | 34 (2) | 53 (3.2) | 87 (5.2) | <0.001 |
| Discharged Home n (%) | 507 (30.5) | 44 (2.7) | 551 (33.2) | <0.001 |
| Admitted to Rehabilitation n (%) | 388 (23.4) | 187 (11.3) | 575 (34.6) |  |
| Transferred to long-term care n (%) | 17 (1) | 141 (8.5) | 158 (9.5) |  |
| Hospitalization for Other Comorbidities n (%) | 75 (4.5) | 142 (8.6) | 217 (13.1) |  |

\* BMI values available in 1508 patients; \*\* Intraparenchymal Volume, Hemorrhage vol. $\geq 30$ ml, ICH score is available in 1501

At 30 days 1487 (89.6%) of patients were alive and 173 (10.4%) died. Most of the deaths were recorded during the hospitalization (156 (9.4%). The mortality at 90 days increased to 251 (15.1%), predominantly males (202/251, 80.5%). Favorable outcome was seen in 59.5% of patients. Individuals with a favorable outcome were significantly younger, and had lower random glucose levels, lower NIHSS and higher GCS scores as shown in table-1. Patients with favorable outcome were more likely to have smaller ICH volume on the initial imaging and supra-tentorial location. Active smoking and dyslipidemia, for unexplainable reasons, were more common in patients with a favorable outcome.

### ICH Localization and imaging characteristics

The most common location for the ICH was sub-cortical in the basal ganglia or the thalamus (1072/1660; 64.6%). ICH location was hemispheric in 280 (16.9%) patients, cerebellar in 152 (9.1%), brainstem in 114 (6.9%) patients, and primary intraventricular 42 (2.5%) patients. Most patients had small hemorrhages. The volume of the hemorrhage was less than 10 ml in patients 906 (60.4%) patients. The volume of the hematoma was between 10-19.9 ml in 311 (20.7%), 20-29.19 in 159 (10.6%) and more than 30 ml in 125 (8.3%) patients. The risk of early mortality and unfavorable outcome increased with increasing volume of the ICH. For each volume of the ICH, patients with infratentorial ICH were more likely to have a higher mortality at 30 and 90 days as shown in figure 3a and b.

**Figure 3:**
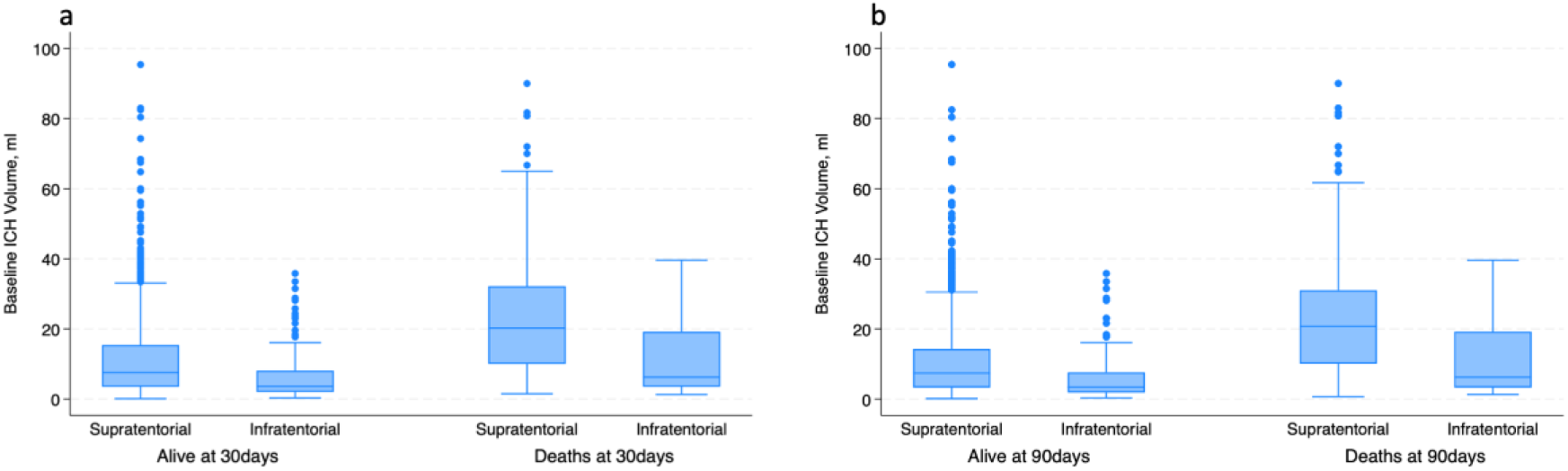
Boxplot. a: shows the distribution of the baseline ICH volume with respect to the location of the ICH and 30-day mortality. Patients with infratentorial locations had higher mortality at lower ICH volume; b: shows the distribution of the baseline ICH volume with respect to the location of the ICH and 90-day mortality. Patients with infratentorial locations had higher mortality at lower ICH volume.

### Comparison of mortality with the ICH-score

As is evident from figure-4, the actual 30-day mortality in our patients was lower at all points of the ICH-score, when compared to the original publication (6). The younger age, smaller volume of ICHs and fewer infratemporal locations likely represent lower 30-day mortality in our study. It is also possible that admission to a stroke unit or ICU in a majority of patients may have also contributed to the lower mortality. Maximally treated ICH patients have been shown to have lower mortality in a previous study (11). There was a proportionate increase in unfavorable outcome with increasing ICH-score as shown in figure-5.

**Figure 4:**
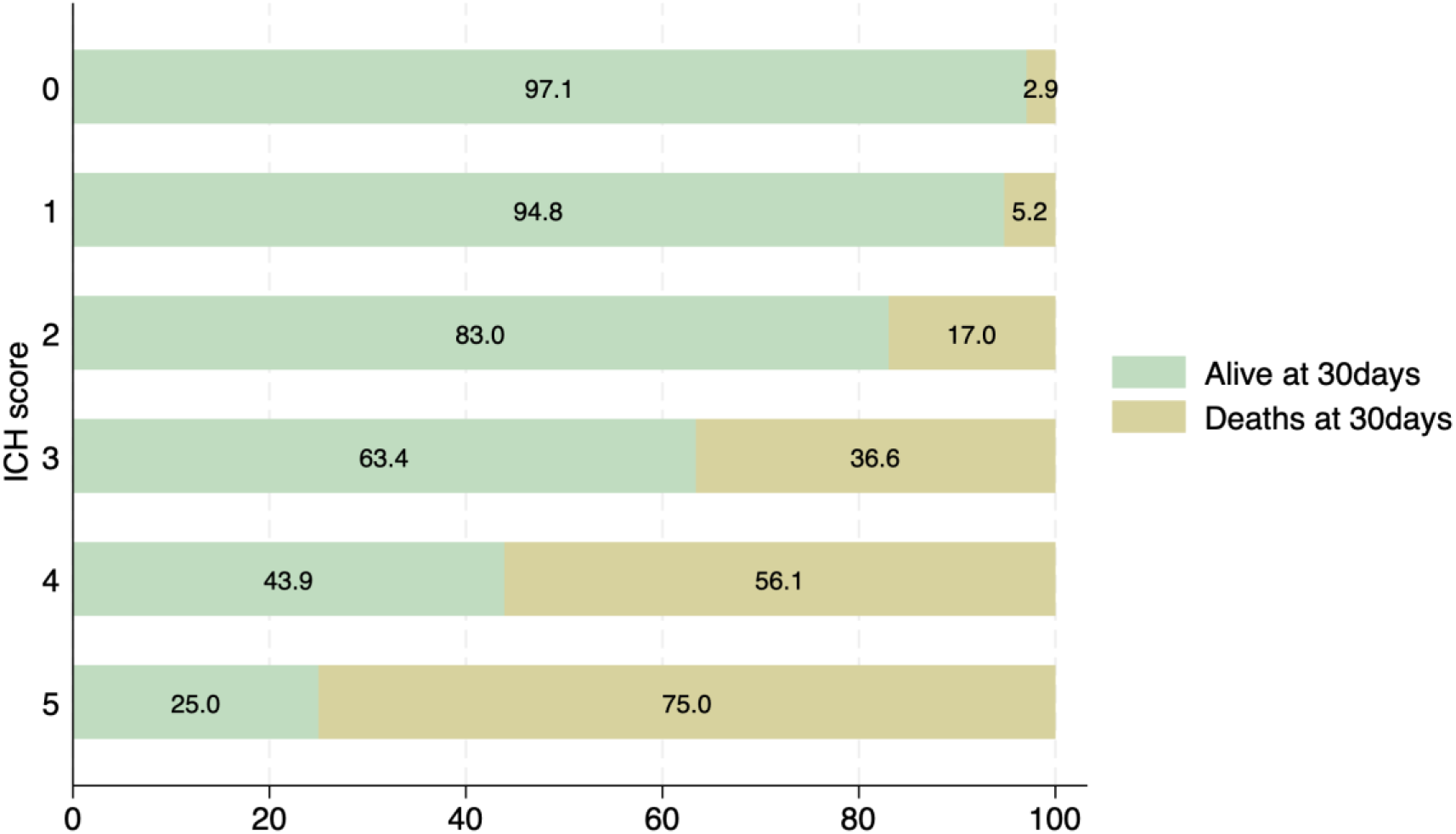
Distribution of patients with ICH score and associated mortality at 30 days. None of the patients had an ICH score of 6.

**Figure 5:**
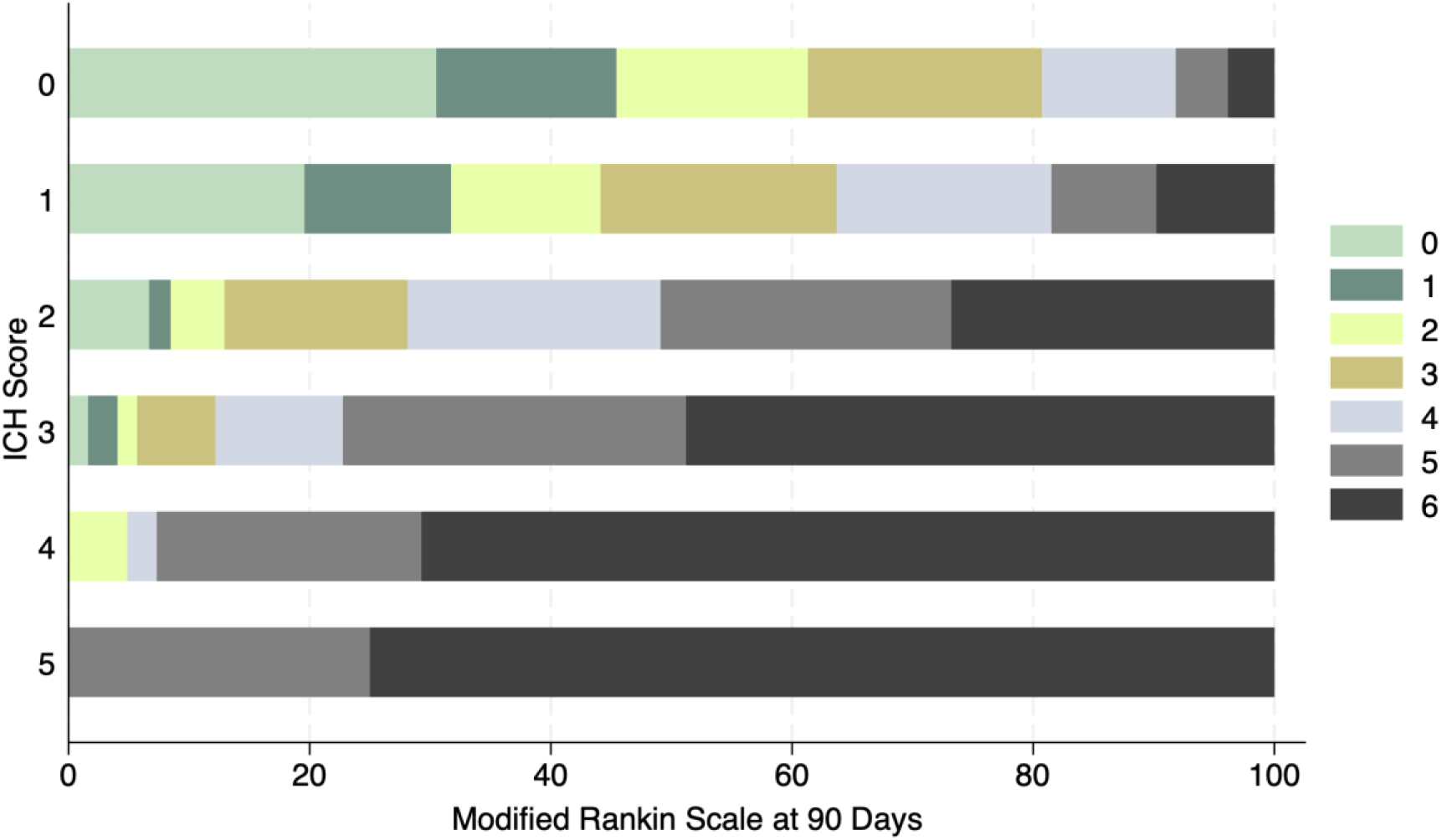
Distribution of mRS at 90 days and ICH score in ICH patients. No patient had a score of 6.

### Antithrombotic medications and ICH prognosis

There were 49 (3%) patients on anticoagulation and 190 (11.4%) on antiplatelet medications at the time of the ICH (supplementary tables 1 & 2). These patients were older (61 (55–74) vs 48 (41–56), p<0.001), and were more likely to have diabetes, CAD, atrial fibrillation and prior stroke. The severity of stroke as measured on the GCS and the NIHSS was not significantly different in patients with or without anticoagulants. ICH volume was also not significantly larger in patients on anticoagulation. Hemispheric location of the ICH was more common in patients taking anticoagulation (35.4% vs 16.8%, p<0.001). Despite the similar volume, and as shown in supplementary table-1, the 30-day (22.4% vs 10.1%; p =0.005) and 90-day (28.6% vs 14.7%; p=0.008) mortality was higher in the anticoagulated patients. The percentage of patients with a favorable outcome (mRS 0-3) at 90-days was also lower in patients on anticoagulation (38.8% vs 61.2%. p<0.003).

As shown in supplementary table 2, patients on antiplatelets were older (63.2±12.1 vs 49.3 ±12.1, p=0.001), and were more likely to have diabetes, CAD, atrial fibrillation and prior stroke. The severity of stroke was not significantly different in patients with or without antiplatelets. ICH volume was also similar in patients on antiplatelets. Hemispheric location of the ICH was more common in patients taking antiplatelets (25.1% vs 16.3%, p <0.001). Despite the similar volume of the ICH, and as shown in supplementary table 2, the 30-day (14.7% vs 9.9%; p=0.04) mortality was higher in patients with antiplatelets. The percentage of patients with a favorable outcome (mRS 0-3) at 90-days was also lower in patients on antiplatelet medications (44.7% vs 55.3%. p<0.001).

### Diabetes and ICH prognosis

There were 564 (34%) patients with known diabetes at the time of the ICH. As supplementary table 3, these patients were older (56.6±13.0 vs 47.1 ±12.4, p< 0.001), and were more likely to have prior stroke history, CAD and atrial fibrillation. The severity of stroke was higher in patients with diabetes. A GCS of 13-15 was seen in 34.4% of patients with diabetes and 47.8% of patients without diabetes (p<0.001). Fewer patients with diabetes had an ICH volume <10 ml (56.4 vs 60.6, p<0.001) and hemispheric location of the ICH was more common in patients with diabetes 19.8% vs 16.6%, p =0.02). The 30-day (13.2% vs 8.8%; p<0.001) and 90-day mortality (18.8% vs 13.6%; p<0.001) was significantly higher in the diabetic patients.

Multivariate analysis of factors that led to an increase in 30-day and 90-day mortality and an unfavorable outcome at 90 days.

The multivariable analysis for the 30-day, 90-day mortality and 90-days unfavorable outcome are shown in table-2. Baseline lower GCS, baseline ICH volume, and infratentorial location, presence of any intraventricular hemorrhage, no surgical intervention was all associated with a higher risk of 30- day and 90-day mortality and higher 90-day unfavorable outcome. The receiver operator curve analysis showed lower GCS score had the highest area-under the curve 0.80, 0.81 and 0.79 among all the variables to predict 30-day, 90-day mortality and 90-day unfavorable outcome (table-3). Baseline ICH volume showed moderate concordance with AUC of 0.69, 0.71 and 0.71 to predict 30- day, 90-day mortality and 90-day unfavorable outcome. Other variables were fair.

**Table 2:** Multivariable Association of Predictors with 30- and 90-day mortality.

| Factor | 30 Days Mortality |  |  | 90 Days Mortality |  |  |
| --- | --- | --- | --- | --- | --- | --- |
|  | uOR (95% CI) | aOR (95% CI) | AUC | uOR (95% CI) | aOR (95% CI) | AUC |
| Age, years | 1.00 (0.99-1.01) | 1.00 (0.99-1.02) | 0.51 | 1.01 (0.99-1.02) | 0.99 (0.98-1.02) | 0.52 |
| Sex | 0.78 (0.53-1.16) | 0.92 (0.58-1.46) | 0.48 | 0.81 (0.58-1.14) | 0.76 (0.45-1.26) | 0.49 |
| Expatriate South Asian/<br>Southeastern Asian status | 0.88 (0.62-1.24) | 1.16 (0.74-1.81) | 0.52 | 0.77 (0.57-1.03) | 0.94 (0.56-1.58) | 0.52 |
| GCS score | 0.76 (0.73-0.79) | 0.78 (0.75-0.82) | 0.80 | 0.74 (0.72-0.77) | 0.79 (0.75-0.83) | 0.81 |
| Baseline Blood Glucose,<br>mg% | 1.08 (1.04-1.12) | 1.02 (0.98-1.06) | 0.62 | 1.08 (1.05-1.11) | 1.02 (0.97-1.07) | 0.64 |
| Use of Antiplatelet<br>Medication | 1.58 (1.02-2.44) | 1.56 (0.92-2.63) | 0.53 | 1.42 (0.96-2.09) | 2.02 (1.13-3.59) | 0.52 |
| ICH Volume, ml | 1.04 (1.03-1.05) | 1.02 (1.01-1.03) | 0.69 | 1.05 (1.04-1.06) | 1.02 (1.01-1.03) | 0.71 |
| Infratentorial Location | 2.82 (1.98-4.01) | 1.90 (1.19-3.02) | 0.57 | 2.72 (1.99-3.71) | 2.40 (1.46-3.94) | 0.56 |
| Any Intraventricular | 2.98 (2.16-4.11) | 1.46 (1.02-2.09) | 0.64 | 2.97 (2.26-3.91) | 1.57 (1.04-2.35) | 0.64 |
| Admission to Intensive<br>care unit | 3.00 (2.11-4.26) | 1.25 (0.82-1.91) | 0.63 | 3.92 (2.87-5.35) | 0.82 (0.51-1.33) | 0.66 |
| Any Surgical Intervention | 0.86 (0.26-2.83) | 0.27 (0.14-0.54) | 0.49 | 0.77 (0.27-2.21) | 0.26 (0.15-0.45) | 0.50 |

uOR, unadjusted Odds Ratio; aOR, adjusted Odds ratio

**Table 3:** Multivariable Association of Predictors with 90-day modified Rankin scale.

| Factor | 90-Day Unfavorable Outcome |  |  |
| --- | --- | --- | --- |
|  | uOR (95% CI) | aOR (95% CI) | AUC |
| Age, years | 1.02 (1.01-1.03) | 1.02 (1.00-1.03) | 0.54 |
| Sex | 0.73 (0.56-0.94) | 0.86 (0.60-1.21) | 0.48 |
| Expatriate South Asian/Southeastern Asian status | 0.68 (0.55-0.85) | 0.99 (0.71-1.39) | 0.53 |
| GCS score | 0.71 (0.68-0.74) | 0.77 (0.73-0.80) | 0.79 |
| Baseline Blood Glucose, mg% | 1.09 (1.06-1.12) | 1.01 (0.99-1.05) | 0.63 |
| Use of Antiplatelet Medication | 1.96 (1.44-2.66) | 1.82 (1.19-2.77) | 0.53 |
| ICH Volume, ml | 1.07 (1.06-1.09) | 1.03 (1.02-1.04) | 0.71 |
| Infratentorial Location | 1.76 (1.35-2.29) | 1.31 (0.89-1.96) | 0.52 |
| Any Intraventricular | 3.02 (2.45-3.72) | 1.57 (1.19-2.08) | 0.63 |
| Admission to the Intensive care unit | 4.49 (3.62-5.54) | 1.69 (1.27-2.26) | 0.68 |
| Any Surgical Intervention | 2.62 (1.28-5.35) | 0.92 (0.58-1.44) | 0.56 |

uOR, unadjusted Odds Ratio; aOR, adjusted Odds ratio

## Discussion

Intracerebral hemorrhage comprised 14.8 % of consecutive stroke patients admitted with acute stroke in Qatar. The patients were young, had male predominance and from multiple ethnicities, reflecting the population dynamics in Qatar where ∼85% of the population is expatriate and mostly comprised of young male workers (26–30). The mortality at 30 days (10.4%) and 90 days (15.1%) was lower in our patients when compared to the previous reports of 30-55% in the literature (2–5). Smaller volume of the hematoma in most of our subjects (only 125 (8.3%) of patients with ICH of ≥ 30.0 ml/cm3)), and smaller number of patients with low GCS of 3-4 [140 (8.4%)] likely contributed to the lower mortality. A recent study in 2023 from Montreal reported a better 30- and 90-day mortality of 16% and 22% respectively from patients admitted to their hospital between 2010-2017 (31). These numbers are comparable to our study and likely reflects improvement in patient care. In our study, 90-day unfavorable outcome (mRS 4-6) was seen in 40.5% patients. Increasing age, presence of vascular risk factors, especially diabetes, and the use of antithrombotic medications, especially anticoagulants, were most likely to associated with an unfavorable outcome.

Most studies on ICH focus on mortality, especially early deaths in the hospital and within the initial 30 days. Patients who survive the initial insult are however left with significant disability. The mRS is used globally to determine recovery following ischemic stroke and has been used in occasional ICH studies to evaluate recovery at 90 days to a year (21,23,32,33,). An mRS of 0-3 (favorable outcome) at 90 days following the ICH was evident in 59.5% of patients in our study. In a smaller study of 243 patients, only 51% of patients reached an mRS of 0-2 at 3 months follow-up and 56% by one-year follow-up (32). The larger study with 3255 patients from China also showed an mRS of 0-2% in 49% of patients at 90- days and 53% by one year (23). In the Swiss Stroke Registry, 2,650 patients with ICH were followed for functional recovery and 33.2% showed good functional recovery (mRS 0-2) at 3 months (33). In a study of 919 patients from the CLEAR-III and MISTIE-III trials, 11.5 % died within 30 days and an mRS of 0-3 was seen in only 14.7% at 30 days (34). Finally, in the recent study from Montreal, similar to our study, favorable outcome (an mRS of 0-3 at 3 months) was evident in 50 percent of patients (31). The mRS at 90-days may therefore be an important measure of recovery in addition to the mortality at 30-days following an ICH to determine prognosis.

An important observation in our series was that the lower rates of death when compared to prior reports (2–5). At 30 days following the ICH, only 10.4 percent of the patients were dead. Lower death rates related to ICH were reported in the recent study from Montreal [30-day mortality of 12%] (31), and the sub-groups of CLEAR-II and MISTIE-III trials (34). The lower mortality rates most likely reflect better hospital care as no new ICH therapies have recently been added to patient care. Similar to ischemic stroke care, stroke unit care also benefits ICH patients (11) and likely contributed to the lower mortality. Other factors that contributed to lower mortality in our patients likely include the younger age of the patients and the smaller size of the hematoma. Overall, 59.5% of patients in our series had a favorable outcome at 90 days. This compares well with the 50% favorable outcome reported at 3.1 months in the series recently published from Montreal (31).

ICH related to anticoagulation was seen in 49 (3%) of our patients. Patients on AC did not have higher NIHSS or lower GCS on admission and the ICH were surprisingly also not larger but more frequently cortical. The mortality and favorable outcome were however higher when compared to patients with no anticoagulation. ICH in patients taking anticoagulant meditations is associated with a significantly higher mortality. In a study from Ontario, Canada the prognosis of ICH in patients with anticoagulation was compared to patients not on anticoagulation. Overall, the in-hospital and one-year mortality was 45.4% and 32.4% respectively (35). There were 149 (11.4%) patients in our series on antiplatelet medications at the time of the ICH. In a recent study of 457 consecutive patients with ICH, 20.5% were on antiplatelet medications. Similar to our results, there were no differences in the in-hospital mortality or 90-day favorable outcome in the two groups (36). In another recent study, prior use of antiplatelet medications was associated with higher risk of hypertension with ICH leading to an increase in mortality and higher rates of unfavourable outcomes (37).

Pre-existing diabetes has previously been shown to adversely affect outcomes in patients with ICH (38). Thirty-four percent of patients in our series had diabetes. Diabetes was associated with more severe symptoms, larger ICHs and higher mortality. A recent meta-analysis of 19 studies documented a modest increase in poor outcome from ICH in patients with diabetes (38). The reasons for the unfavorable outcome and higher mortality are not clear but may include increased disruption of the blood-brain-barrier leading to more severe edema formation, increased activation of neuroinflammatory mechanisms and cognitive impairment that may slow recovery efforts (39).

In our study, we also compare outcome in ICH from multiple ethnicities, including Arabs, South East Asians, Philippians and Africans. There are a few studies e documenting outcomes in ICH from ethnic communities. These include studies on the clinical course of ICH from Qatar (16,17) Pakistan (18), Africa (19), Saudi Arabia (20), Hong Kong (21), Philippine (22), China (23), India (24), and Singapore (25). The number of patients in most such series is small (19–22) or the information collected retrospectively (18,21,22,25). Similar to our study, the age at presentation was 58 years from India (24), 57 years from Pakistan (18), 53.7 years from Congo (19), 60. 3 years from Saudi Arabia (20) and 62 years from Chana (23). In most series there was a predominance of men and the reported mortality was between 20% (23) and 50% (24). The volume of ICH was mostly less than 25 ml/cm although was 49.9 in the small series of 60 patients from South India (24). The larger ICH volume in the small study from India may account for the higher 50% mortality in the study (24).

There are some limitations to our research. Although the patients were all entered into the database prospectively, this is a retrospective analysis of the registry. Secondly, the analysis is from a single center in one country. The comparison of clinical and outcome data in a large number of patients from multiple ethnicities is, we believe, a major strength of the study. Thirdly, our follow-up is limited to the 90-day outcome as part of the registry data collection. As shown in the study by Hemphill from USA (6) and Wang et al from China (23), recovery continues well beyond the initial 90 days, we do not have long-term outcome information from our research.

In summary, our study outlines the clinical and radiological features of ICH in a large multiethnic cohort of patients from a single center in Qatar. Our patients were younger and the hematomas were most frequently seen in the sub-cortical thalamic and basal ganglion region. The 30-day mortality was significantly better than what has been previously reported. The lower mortality may be related to the younger age at presentation, fewer patients with sub-tentorial hemorrhages and the small volume of the hematomas. Despite the lower mortality at 30 days, favorable outcome was evident in only 44.5% patients at 90 days.

## Ethical approval

The study was approved by the Institutional Review Board, Hamad Medical Corporation at the Medical Research Centre (MRC-01-18-102)

## Guarantor

Dr. Ashfaq Shuaib, also the corresponding author.

## Data availability statement

No additional data available

## Informed consent

Not applicable

## Contributorship

### Concept, design, and draft

Naveed Akhtar, Mahesh Kate and Ashfaq Shuaib

### Acquisition, analysis, interpretation of data, technical and administrative support

Naveed, Blessy, Deborah, Sujatha, Ryan, Shobana, Naveed Akhtar, Ashfaq Shuaib

### Critical review

Saadat Kamran, Ashfaq Shuaib

### Statistical analysis

Naveed Akhtar

